# Elevated Calcium-Phosphate Product (CPP) in Chronic Kidney Disease (CKD): A Silent Predictor of Poor PCI (Percutaneous Coronary Intervention) Outcomes: A Propensity Score Matched Analysis

**DOI:** 10.64898/2026.03.05.26347359

**Authors:** Muhammad Raffey Shabbir, Waseh Ahsan, Meenal Sikander, Ahmad Baig, Syed Muhammed Salman Hassan, Abdul Manaf, Syeda Aleezey Jibran, Mishal Zehra, Nooria Saif, Uzair Majeed, Salman Khalid, Naeem Tahirkheli

**Author notes:** **Corresponding author** Muhammad Raffey Shabbir, MD | | Marshfield Clinic, Sanford Health, Marshfield, Wisconsin, USA. **Statements and Declarations**. **Author Approval:** All authors have seen and approved the manuscript. **Ethical Approval:** No ethical approval was required for the study. **Consent:** No consent was needed. **Data Availability Statement:** All data generated or analyzed during this study are included in this article. Further inquiries can be directed at the corresponding author.

## Abstract

**Background:** An elevated calcium-phosphate product (CPP), defined as a product of serum calcium and serum phosphate, is a hallmark of CKD-mineral and bone disorder and has been implicated in accelerated coronary artery calcification, arterial stiffness, and left ventricular hypertrophy. These pathological changes contribute to adverse cardiovascular outcomes. While prior studies have shown worse percutaneous coronary outcomes (PCI) outcomes in CKD patients overall, the prognostic impact of CPP levels remains underexplored. The objective is to evaluate post-PCI outcomes in CKD patients with and without hypercalcemia and hyperphosphatemia.

**Methods:** A retrospective cohort analysis was conducted using the TriNetX U.S. Collaborative Network, focusing on adult patients with CKD undergoing PCI. Patients were grouped based on serum calcium and phosphorus levels, with those having hypercalcemia and hyperphosphatemia compared to those without. Diagnoses and procedures were identified using ICD-10 and CPT codes. Propensity score matching was applied to account for differences between groups. Post PCI outcomes were analyzed. Primary outcome was all-cause mortality. Secondary outcomes encompass coronary artery bypass grafting (CABG), myocardial infarction (MI), in-stent re-stenosis [redo PCI] and target vessel revascularization, heart failure (HF) exacerbations, and peri-/ post-procedural complications were assessed within a 5-year follow-up period. Kaplan-Meier analysis with log-rank was used for statistical comparisons, with significance set at p<0.05.

**Results:** The elevated CPP group was significantly associated with increased post-PCI all-cause mortality [hazard ratio (HR) 1.428], in-stent restenosis [HR 1.589], heart failure exacerbations [HR 1.492], and recurrent angina or MI [HR 1.396]. No significant differences were found in rates of post PCI CABG, periprocedural complications (postprocedural cardiac insufficiency, postprocedural cardiac arrest, postprocedural heart failure, intraoperative cerebrovascular infarction, postprocedural cerebrovascular infarction, and intraoperative cardiac arrest), or redo PCI.

**Conclusion:** In this propensity score-matched analysis, elevated CPP in CKD patients undergoing PCI was independently associated with worse outcomes, including higher mortality and cardiovascular event rates. These findings highlight the prognostic value of CPP and the need for closer metabolic monitoring and individualized risk stratification.

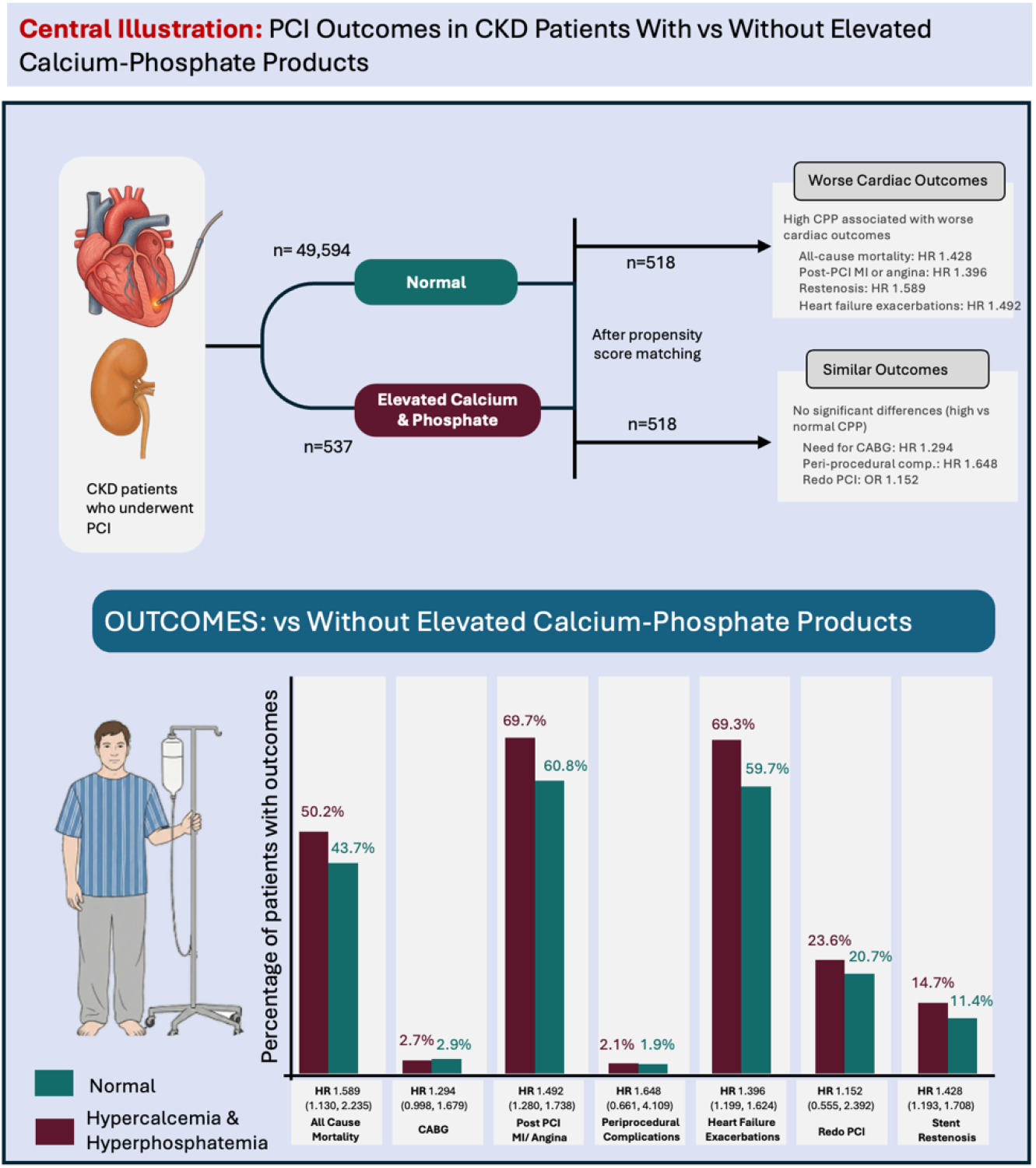

## Introduction

Chronic kidney disease (CKD) is among the most prevalent chronic conditions and is associated with substantial cardiovascular morbidity and mortality. The burden of CKD continues to rise, with data from the Centers for Disease Control and Prevention indicating that approximately 1 in 7 adults—nearly 35.5 million individuals—in the United States are affected by this condition (1). Cardiovascular disease remains a leading cause of death among patients with CKD (2). Notably, cardiovascular risk is evident even in the initial stages of CKD and increases progressively with advancing renal dysfunction (3).

PCI is a widely utilized revascularization strategy for the management of acute myocardial infarction (MI). However, patients with CKD undergoing PCI experience a higher incidence of peri-procedural and long-term complications compared with those with preserved renal function (4).

The kidneys play a critical role in maintaining calcium–phosphate homeostasis. In CKD, disruption of these regulatory mechanisms leads to variations in serum calcium and phosphate levels. The calcium–phosphate product (CPP), calculated as the product of serum calcium and phosphate concentrations, is a key biomarker reflecting this metabolic derangement. (5) Elevated CPP has been associated with vascular calcification and an increased risk of adverse cardiovascular events in patients with CKD (6).

In this study, we independently evaluated the association between elevated pre-procedural CPP levels and clinical outcomes following PCI.

## Methods

### Data Availability

All data generated or analyzed during this study are included in this article and its supplementary materials. Further inquiries can be directed at the corresponding author.

### Data Source and Study Design

We conducted a retrospective cohort study using the TriNetX, LLC platform, a global federated health research network that provides access to de-identified electronic health records (EHRs) from participating healthcare organizations (HCOs). TriNetX aggregates data across diverse care settings, including hospitals, primary care clinics, and specialty practices, and includes records from both insured and uninsured patients. The available data included information about the demographics, diagnoses (based on the International Classification of Diseases, Tenth Revision, Clinical Modification, ICD-10-CM codes), procedures (coded in The International Classification of Diseases, Tenth Revision, Procedure Coding System, ICD-10-PCS or Current Procedural Terminology, CPT), medication (coded in Veterans Affairs National Formulary), laboratory tests (coded in Logical Observation Identifiers Names and Codes, LOINC), genomics (coded in Human Genome Variation Society, HGVS), and healthcare utilization.

Our study utilized the TriNetX US Collaborative Network, which included 47 healthcare organizations (HCOs) and over 76 million de-identified patients at the time of data extraction in 2025. The platform employs natural language processing (NLP) to improve the structured data fields.

### Ethics Statement

The TriNetX platform is compliant with the Health Insurance Portability & Accountability Act and General Data Protection Regulation (GDPR). The Western Institutional Review Board (WIRB) has granted a waiver of informed consent for studies utilizing TriNetX data. Ethical approval was deemed unnecessary, consistent with Section §164.514 of the HIPAA Privacy Rule.

### Data Quality

TriNetX ensures data quality through routine evaluations of conformance, completeness, and plausibility, utilizing both internal and external validation methods. These quality assurance processes support its utility in real-world evidence generation and randomized controlled trial (RCT) planning.

### Cohort Definition and Eligibility

Cohort construction followed a two-step process: 1) Defining eligibility criteria using structured query parameters 2) Executing built-in analysis tools within the TriNetX platform.

Adults aged 40-90, male or female, were identified and included in the study. Patients with pre-existing chronic kidney disease (ICD-10-N 18) who underwent PCI (CPT 1021163, 1021164, 1021165, 1021166, 1021166, 1021167, and 1021168) prior to June 1, 2020, were made part of the study.

Utilizing US collaborative network, ICD-10 codes were utilized for defining chronic kidney disease (N18), hyperphosphatemia (E83.30 & E83.39) and hypercalcemia (E83.52). CPP was calculated as the product of serum calcium (mg/dL) and serum phosphate (mg/dL), expressed in mg^2^/dL^2^.

Additionally, PCI was included using CPT Codes (1021163, 1021164, 1021165, 1021166, 1021167, and 1021168). Patients were divided into two separate cohorts, depending on whether they had hypercalcemia and hyperphosphatemia leading to relatively higher calcium phosphate product (CPP) at the time of PCI or not. Using date constraint event relationship, in elevated CPP group any instance of PCI occurred within 1 month or any time after documented hypercalcemia and hyperphosphatemia.

Using same ICD and CPT codes, second cohort incorporated patients with CKD who never had a diagnosis of hypercalcemia or hyperphosphatemia leading to lower CPP and underwent PCI. This approach created clear timing between exposures and ensured comparability in the two groups. Patients with primary hyperparathyroidism (E21.0), functional disorders of polymorphonuclear neutrophils (D71), hypervitaminosis (E67.3) were excluded to avoid confounding causes of altered calcium or phosphate homeostasis.

### Propensity Scoring and Matching

To avoid immortal time bias, we defined the index event as the date when each patient met all the inclusion criteria. To mimic a randomized study and reduce confounding, we used a 1:1 propensity scores matching method. This involved a greedy nearest neighbor algorithm without replacement, with a caliper width of 0.1 times the pooled standard deviation of the logit of the propensity score. Patients who couldn’t’t be matched were removed from the analysis.

A total of 26 baseline covariates are used in matching process; baseline demographics (age, sex, race, and ethnicity), diagnoses (hypertensive diseases; disorders of lipoprotein metabolism and other lipidemia; diabetes mellitus; end-stage renal disease; and chronic kidney disease stages (1, 2, 3, 4, and 5), medication use (beta blockers, anti-lipid agents, antianginals, platelet aggregation inhibitors), and BMI (category indicators for 25–29.9 kg/m^2^, 30–34.9 kg/m^2^, 35– 39.9 kg/m^2^, and greater than 40 kg/m^2^), consistent with clinically relevant covariates that could influence calcium-phosphate balance or cardiovascular outcomes. Covariate balance was assessed using standardized mean differences (SMDs), with an SMD < 0.1 indicating acceptable balance.

### Outcomes and end points

The study outcomes were assessed within a five-year follow-up period following the index PCI. Outcomes were identified using validated CPT and ICD-10 codes, including all-cause mortality, coronary artery bypass graft (1006199, 1006207, 1006216), myocardial infarction (I20 -I22), stent restenosis (T82.855), heart failure exacerbation ((I50.2–I50.4) and peri/post procedural complications including cardiac insufficiency, cardiac arrest, heart failure, or cerebrovascular infarction (I97.11, I97.12, I97.13, I97.81, I97.85, I97.71, I97.790).

### Statistical Methods

All statistical analyses were performed using the built-in analytic tools of the TriNetX platform. Continuous variables were presented as means ± standard deviations and compared using independent samples t-tests, while categorical variables were summarized as counts and percentages and compared using Chi-square tests. Survival outcomes were analyzed using the Kaplan-Meier method, and survival distributions between groups were compared using the log-rank test. Hazard ratios (HRs) with 95% confidence intervals (CIs) were calculated to assess the relative risk of outcomes. The log-rank test statistic was interpreted using a chi-square distribution with 1 degree of freedom (df = 1), as appropriate for two-group comparisons.

To estimate measures of association between the matched cohorts, we used risk ratios (RRs), odds ratios (ORs), and absolute risk differences (RDs), each reported with corresponding 95% CIs. These were calculated directly within the platform’s outcome comparison tools. A two-sided p-value < 0.05 was considered statistically significant. All analyses were conducted entirely within the TriNetX platform; no data was exported for external statistical analysis.

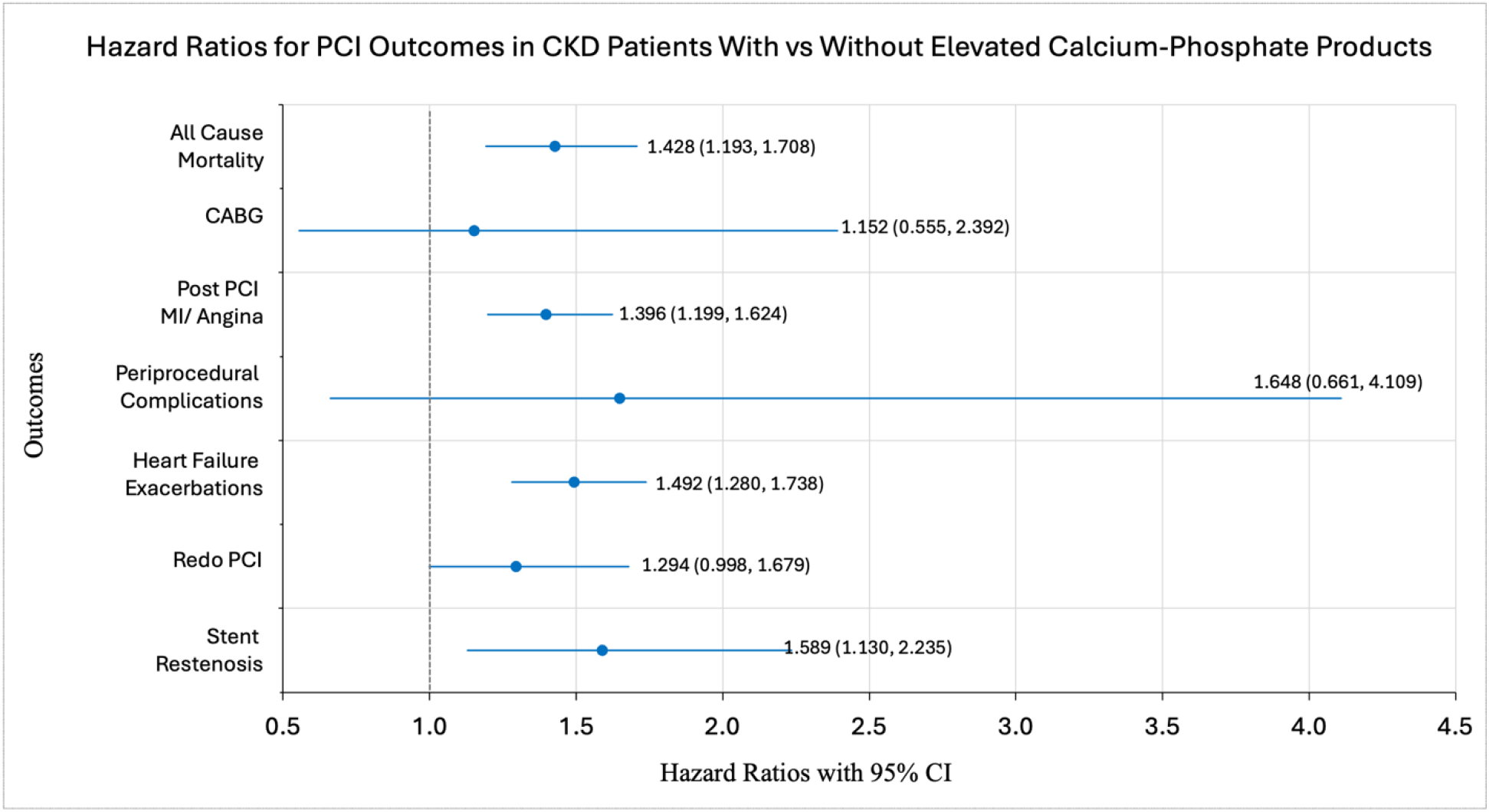

## Results

### Study population

Our study compares the outcomes of PCI in CKD patients with high calcium phosphate product (CPP) levels to those with normal CPP levels. CPP was calculated as the product of serum calcium (mg/dL) and serum phosphate (mg/dL), expressed in mg^2^/dL^2^.

Patients were stratified based on the presence of diagnostic codes for hypercalcemia and hyperphosphatemia as identified within the TriNetX database. Individuals meeting criteria for these metabolic abnormalities comprised the higher CPP group, whereas those without such diagnostic codes comprised the lower CPP group. The mean CPP in the hypercalcemia/hyperphosphatemia group was 39.1 mg^2^/dL^2^, compared with 32.45 mg^2^/dL^2^ in the comparison group.

A total of 50,131 patients with CKD underwent PCI. At the time of PCI, 49,594 had lower CPP, while 537 patients had higher CPP.

Prior to propensity score matching, there were significant differences in baseline demographics, comorbidities, medication use, and laboratory parameters between the two groups. Following 1:1 propensity score matching, 518 patients remained in each cohort. Matching achieved excellent covariate balance, with the standardized mean differences reduced to <0.05 for nearly all variables, and no statistically significant differences observed between groups in demographics, comorbidities, medication use, or laboratory values. (Table 1. Basic Characteristics Table)

**Table 1.** Baseline characteristics following propensity matching. No statistically significant differences observed between the two groups (p>0.05). Abbreviations: Std diff: Standard difference, BMI: Body Mass Index.

| Variables | Hypercalcemia and hyperphosphatemia | Excluding Hypercalcemia and hyperphosphatemia | P-Value | Std diff. |
| --- | --- | --- | --- | --- |
| **Demographics** |  |  |  |  |
| Current Age | 70.5 +/- 10.9 | 71.1 +/- 11.5 | 0.367 | 0.056 |
| White | 324 | 315 | 0.565 | 0.036 |
| Female | 216 | 230 | 0.380 | 0.055 |
| Non-Hispanic or Latino | 389 | 400 | 0.423 | 0.050 |
| Hispanic or Latino | 47 | 36 | 0.208 | 0.078 |
| Black or African American | 126 | 135 | 0.520 | 0.040 |
| Male | 283 | 271 | 0.455 | 0.046 |

| **Diagnosis** |  |  |  |  |
| --- | --- | --- | --- | --- |
| Hypertensive diseases | 511 | 509 | 0.614 | 0.031 |
| Disorders of lipoprotein metabolism and other lipidemia | 477 | 470 | 0.438 | 0.048 |
| Diabetes mellitus | 417 | 431 | 0.259 | 0.070 |
| End stage renal disease | 374 | 383 | 0.528 | 0.039 |
| Chronic kidney disease, stage 3 (moderate) | 272 | 282 | 0.533 | 0.039 |
| Chronic kidney disease, stage 5 | 244 | 236 | 0.618 | 0.031 |
| Chronic kidney disease, stage 4 (severe) | 241 | 241 | 1 | <0.001 |
| Chronic kidney disease, stage 2 (mild) | 80 | 85 | 0.671 | 0.026 |
| Chronic kidney disease, stage 1 | 30 | 31 | 0.895 | 0.008 |
| **Medication** |  |  |  |  |
| BETA BLOCKERS/RELATED | 486 | 480 | 0.458 | 0.046 |
| ANTILIPEMIC AGENTS | 481 | 470 | 0.213 | 0.077 |
| ANTIANGINALS | 401 | 388 | 0.343 | 0.059 |
| PLATELET AGGREGATION INHIBITORS | 493 | 495 | 0.768 | 0.018 |
| ANTICOAGULANTS | 480 | 476 | 0.642 | 0.029 |

Table 1: Baseline characteristics following propensity matching. No statistically significant differences observed between the two groups ( $p>0.05$ ).
| Laboratory |  |  |  |  |
| --- | --- | --- | --- | --- |
| BMI | 29.0 +/- 6.4 | 30.8 +/- 6.9 | <0.001 | 0.277 |
| 25 - 29.90 kg/m2 | 317 | 294 | 0.146 | 0.090 |
| 30 - 34.90 kg/m2 | 278 | 281 | 0.852 | 0.012 |
| 35 - 39.90 kg/m2 | 173 | 169 | 0.792 | 0.016 |
| >40 kg/m2 | 111 | 121 | 0.456 | 0.046 |

### Primary outcome

Over a follow-up of 5 years after PCI, elevated CPP exhibited significantly higher hazard ratios (HR) for various adverse outcomes. (Table 2: Outcomes Table)

**Table 2.** Outcomes table comparing post-PCI outcomes between Elevated Calcium-Phosphate Product vs Normal Calcium-Phosphate groups. Product Hazard Ratio (HR), Risk ratio (RR), Odds Ratio (OR), and Risk difference (RD) for outcomes following PCI. Abbreviations/key: HR=Hazard Ratio, CI=Confidence Interval; RD=Risk Difference; RR=Risk Ratio; OR=Odds ratio

| Name | Hazard Ratio<br>HR (95%CI) | Kaplan-Meier<br>log-rank test |  |  | Risk ratio<br>RR (95% CI) | Odds Ratio<br>OR (95% CI) | Risk Difference<br>RD (95% CI) |
| --- | --- | --- | --- | --- | --- | --- | --- |
| | | $\chi^2$ | df | p | | | |
| All-Cause Mortality | 1.428<br>(1.193, 1.708) | 15.2 | 1 | <0.05 | 1.149<br>(1.008, 1.309) | 1.299<br>(1.016, 1.660) | 0.065<br>(0.004, 0.126) |
| CABG | 1.152<br>(0.555, 2.392) | 0.1 | 1 | 0.704 | 0.933<br>(0.455, 1.914) | 0.931<br>(0.445, 1.950) | -0.002<br>(-0.022, 0.018) |
| Post PCI<br>MI or Angina | 1.396<br>(1.199, 1.624) | 19.2 | 1 | <0.05 | 1.146<br>(1.048, 1.253) | 1.482<br>(1.146, 1.917) | 0.089<br>(0.031, 0.147) |
| Periprocedural<br>Complications | 1.648<br>(0.661, 4.109) | 1.1 | 1 | 0.279 | 1.1<br>(0.471, 2.568) | 1.102<br>(0.464, 2.618) | 0.002<br>(-0.015, 0.019) |
| Heart Failure<br>Exacerbations | 1.492<br>(1.280, 1.738) | 26.9 | 1 | <0.05 | 1.162<br>(1.061, 1.273) | 1.527<br>(1.182, 1.973) | 0.097<br>(0.039, 0.155) |
| Redo PCI | 1.294<br>(0.998, 1.679) | 3.8 | 1 | 0.051 | 1.14<br>(0.907, 1.434) | 1.183<br>(0.882, 1.588) | 0.029<br>(-0.022, 0.079) |
| Stent<br>Restenosis | 1.589<br>(1.130, 2.235) | 7.2 | 1 | <0.05 | 1.288<br>(0.938, 1.770) | 1.338<br>(0.929, 1.926) | 0.033<br>(-0.008, 0.074) |

All-cause mortality occurred in 258 individuals with elevated CPP (50.2%) and 225 individuals with normal CPP (43.6%), corresponding to a hazard ratio (HR) of 1.428 (95% CI: 1.193-1.709; p < 0.05).

### Secondary outcomes

In elevated CPP cohort, significantly higher hazard ratios were observed for post PCI myocardial infarction or angina (HR: 1.396; 95% CI: 1.199-1.624; p<0.05), stent restenosis (HR: 1.589; 95% CI:1.130-2.235; p<0.05) and heart failure exacerbations (HR:1.492; 95% CI:1.280-1.738; p<0.05).

No significant difference was seen in CABG (HR=1.152, 95% CI:0.555-2.392; P=0.704), peri-procedural complications (HR=1.648; 95% CI: 0.661-4.109; p=0.279) and redo-PCI (HR=1.294; 95% CI: 0.998-1.679; p=0.051) between both the groups, suggesting minimal association of CPP on these specific outcomes (Table 2)

**Fig 1.**
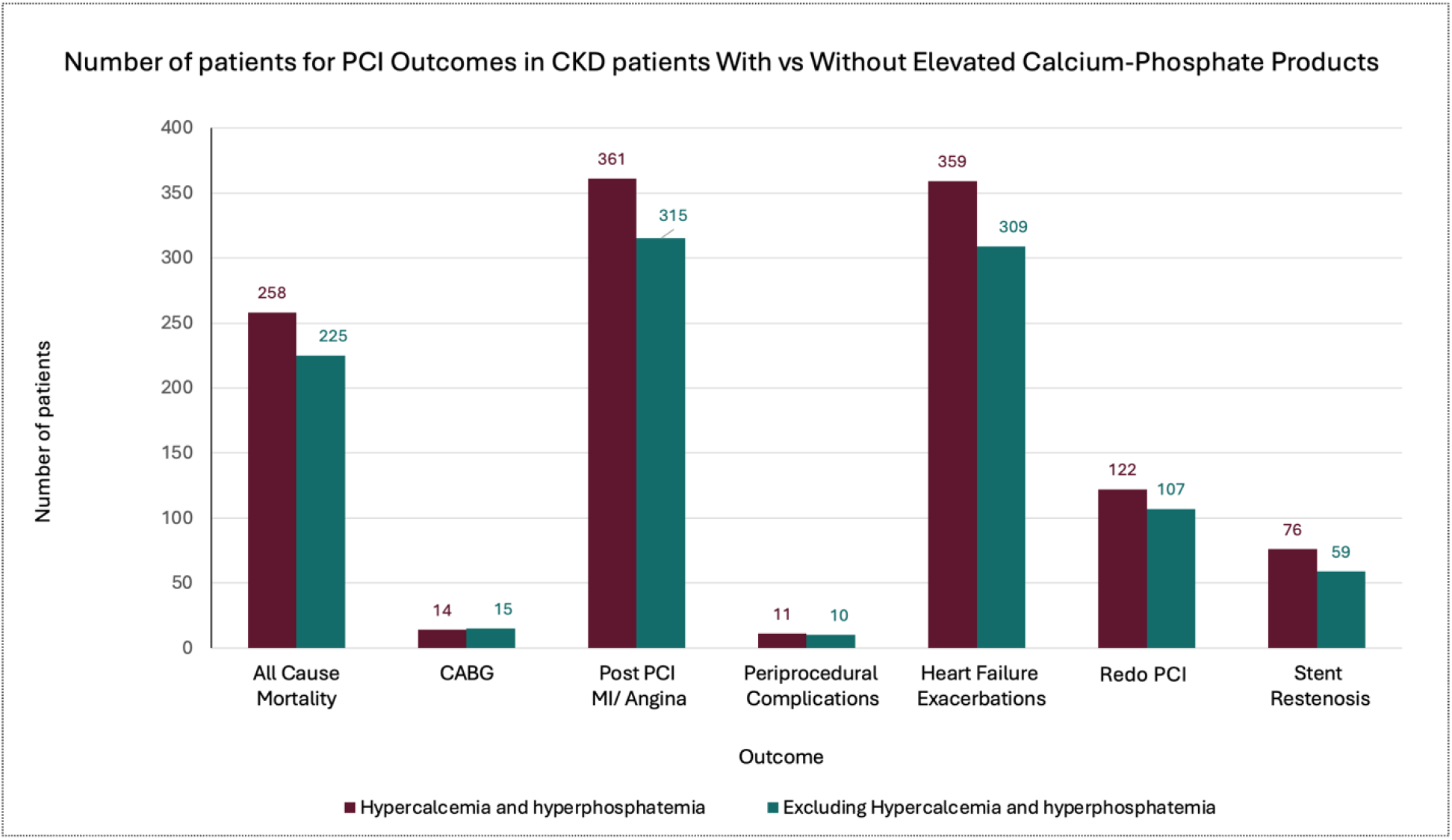
Bar chart comparing the number of outcomes post PCI between patients having high calcium and phosphate vs those having normal.

## Discussion

In this retrospective cohort study using the TriNetX registry, a real-world data source, we evaluated the difference in outcomes of PCI in patients with chronic kidney disease with elevated vs normal calcium phosphate product. The primary outcome of this study was all-cause mortality which was significantly higher in patients with elevated CPP. Several other secondary outcomes included post PCI myocardial infarction/angina, stent restenosis, and heart failure exacerbations were also observed in this group. Clinically, abnormal calcium–phosphate status may identify a higher-risk subgroup of patients with chronic kidney disease undergoing PCI who could benefit from closer post-procedural surveillance and optimization of secondary prevention.

Chronic kidney disease is associated with substantially increased incidence of coronary artery disease. Due to presence of multiple comorbidities, it complicates the procedural and post-procedural outcomes (7). Disturbances in mineral metabolism are common in chronic kidney disease, which leads to elevated levels of calcium phosphate products that increase inflammation, vessel stiffness, and cardiovascular calcifications (8). This process is relevant particularly in PCIs as vascular calcifications and stiffness affect procedural success and stent performance. While there are studies in medical literature about the association of calcium phosphate products and coronary artery disease, limited studies evaluate PCI outcomes stratified by CPP levels based on real-world data. Understanding this relationship may help refine risk stratification in a high-risk population undergoing coronary revascularization.

Prior studies have demonstrated increased risk of cardiovascular morbidity and mortality in patients with CKD. For instance, Go et al observed an independent association between risk of cardiovascular events with reduced GFR (9), while Sarnak et al demonstrated increased risk of cardiovascular mortality in patients with end stage renal disease on dialysis (10). Chronic kidney disease is also characterized by disturbances in mineral metabolism and increased accumulation of calcium–phosphate products. In this context, Block et al, observed independent association of abnormal mineral metabolism in dialysis patients with cardiovascular morbidity and mortality (11). Mutema et al, demonstrated worse outcomes and higher prevalence of comorbidities in patients with CKD undergoing PCI compared to patients without CKD (12). However, these analyses did not stratify outcomes by calcium–phosphate product levels. Our study extends existing literature by leveraging large-scale real-world data to evaluate PCI outcomes stratified by calcium–phosphate product levels within a CKD population, an area in which data remain limited.

Several pathophysiological mechanisms can explain the association between elevated calcium phosphate levels in CKD patients and adverse post-PCI outcomes. Increased levels of calcium-phosphate products in CKD patients are major contributors to vascular calcification and have been associated with increased mortality (13). Calcified, stiff and less pliable coronary vessels may prevent proper stent expansion and healing, thereby increasing the risk of stent restenosis and recurrent ischemic events. Furthermore, arterial stiffness may increase left ventricular afterload leading to heart failure exacerbations and increased mortality following PCI. Disordered mineral metabolism and disposition is also associated with inflammation and abnormal endothelial dysfunction (14). This can promote endothelial proliferation and neointimal hyperplasia leading to stent-restenosis and recurrent ischemic events.

Secondary outcomes in our study further help us understand the clinical significance of elevated CPP levels and their association with increased mortality. We observed higher incidences of post-PCI angina or myocardial infarction in patients with elevated CPP levels. This may reflect recurrent ischemia following revascularization. Higher incidence of stent restenosis reflects compromised durability of PCI in this population. Furthermore, we also observed that the patients with elevated CPP levels experience more heart failure exacerbations which may represent downstream consequences of both increased arterial stiffness and recurrent ischemia, amplifying overall cardiovascular risk. Taking together these secondary outcomes provide a plausible explanation of increased mortality post-PCI in CKD patients with elevated CPP.

This study has several notable strengths and some limitations that warrant consideration. We used a large-scale real-world dataset which reflects a diverse population of patients with CKD undergoing PCI. Using propensity score matching helped balance baseline demographics and clinical characteristics between comparison groups. Nevertheless, several limitations warrant consideration. First, the retrospective nature of the study precludes definitive causal inference. Residual confounding from unmeasured variables remains possible. Second, reliance on ICD and CPT codes may introduce misclassification, as coding variations in documentation practices can affect diagnostic accuracy. Additionally, some of the granular data were unavailable like procedural characteristics, medication adherence, and serial calcium-phosphate product levels. The findings in our study should be interpreted as associative and hypothesis-generating.

Our study highlights the role of CPP as a marker of risk of adverse events in a vulnerable population undergoing coronary revascularization. Incorporating calcium phosphate status in patients with chronic kidney disease undergoing PCI could help stratify patients who could benefit from close post-procedural surveillance and follow-up. Future prospective studies are warranted to highlight CPP as a marker of increased risk of poor post-PCI outcomes in patients with CKD and develop targeted therapies to improve prognosis. Overall, these findings underscore the importance of metabolic factors in cardiovascular risk assessment among patients with CKD.

## Data Availability

All data generated or analyzed during this study are included in this article.

https://live.trinetx.com/

## Acknowledgments

None

